# A novel approach to identify the fingerprint of stroke gait using deep unsupervised learning

**DOI:** 10.1101/2024.12.19.24319338

**Authors:** Sina David, Sonja Georgievska, Cunliang Geng, Yang Liu, Michiel Punt

## Abstract

**Background:** The gait pattern results from a complex interaction of several body parts, orchestrated by the (central) nervous system that controls the active and passive systems of the body. An impairment of gait due to a stroke results in a decline in quality of life and independence. Setting up efficient gait training requires an objective and wholesome assessment of the patient’s movement pattern to target individual gait alterations. However, current assessment tools are limited in their ability to capture the complexity of the movement and the amount of data acquired during gait analysis.

**Aims:** In this study, we explore the potential of variational autoencoders (VAE) to learn and recognise different gait patterns within both, pathologic and healthy gait.

**Methods:** For this purpose, the lower-limb joint angles of 71 participants (29 stroke survivors, 42 healthy controls) were used to train and test a VAE.

**Results:** The good reconstruction results (range r = 0.52 - 0.91, average normalized RMSE 23.36 % ± 4.13) indicate that VAEs extract meaningful information from the gait pattern. Furthermore, the extracted latent features are sensitive enough to distinguish between the gait patterns of stroke survivors and a healthy cohort (p<0.001).

**Conclusions:** The presented approach allows the assessment of gait data in an objective and wholesome manner, thereby integrating the individual characteristics of each person’s gait, making it a suitable tool for monitoring the progress of rehabilitation efforts.

## Introduction

Human gait is highly individual just like a fingerprint. The fact that one can recognize a known person from a far distance is utilized for gait biometrics, identifying the individual solely by their gait pattern [1–3].

A gait pattern results from a complex interaction of several body parts, orchestrated by the (central) nervous system. Therefore, pathologic gait is an expression of altered motor function. Individuals who have had a stroke exhibit different gait patterns than healthy individuals [4]. Their gait pattern is usually not optimal resulting in an increased energy cost [5,6] and a reduction of stability of walking [7] when compared to healthy elderly.

As walking is the most frequently executed form of locomotion, this impacts participation and quality of life [8]. Therefore, regaining gait quality is one of the main goals of stroke rehabilitation [9].

To assess gait quality and potential therapy success, researchers and clinicians seek to identify meaningful information in the gait pattern using instrumented 3D gait analysis to collect predefined segment motions [10]. The result is a high dimensional data set where each time series non-linearly interacts with others resulting in a problem of high complexity.[11] One common solution to reduce complexity is the preselection of variables of interest. However, this selection is dependent on the background and experience which might result in partly subjective interpretations of the formerly objective gait data [12,13]. Moreover, this approach could result in the neglect of key features by focusing on predefined outcome measures and does not reflect the complexity of human gait where the emphasis should be put on the interaction of different variables [11,14].

Lately, advanced data processing and analysis methods were developed to tackle the complexity problem. Both supervised and unsupervised methods showed promise to classify different stroke gait patterns. Algorithms such as support vector machines, principal component analysis, and decision trees have been studied for their ability to extract clinically relevant information from gait data [15–20], distinguish patient [21,22] or identify the representatives of individual gait [23].

These approaches although objective, are often in need of conventional gait features for training and clustering algorithms are not capable of reflecting the individual characteristics of a patient’s gait pattern [14] or neglect that the differences between gait patterns are continuous. To this end, we propose a novel method for analysing 3D gait data. We opt for a wholesome quantitative approach that incorporates all collected information utilizing the interactions between multiple joints and motion planes, extracts characteristics of the input data and therefore objectively reduces the number of dimensions.

Variational autoencoders (VAE) are a type of generative model that can learn a compressed, probabilistic representation of high-dimensional data, such as gait patterns [24]. This representation, known as a latent space, can capture the essential features of the data and enable efficient analysis. A VAE extracts a defined number of latent features by encoding the input data as a Gaussian distribution over the latent space, rather than only as a vector [24]. This allows for regularization of the latent space in addition to minimization of the reconstruction error. Therefore, VAEs are an unsupervised method to generate regular latent representations of the input data [25].

The aim of this paper is twofold. First, we propose a new method based on a VAE to learn gait patterns, assessed via the network’s reconstruction accuracy. Second, we aim to do a first exploration of to what extent the VAE’s latent space can be used to represent an individual’s gait fingerprint. We do so in two ways, (1) by exploring whether the gait pattern of each participant forms clusters distinguishable from another and (2) by analysing whether the location of the participant within the latent space is meaningful, expressed through different gait patterns. Further, we hypothesized that the area covered by a stroke survivor’s data within the latent space is larger than the areas of the healthy controls, due to the higher expected variability contained in stroke gait [26].

## Methods

### Participants

For this study, two data sets were combined. The first sample contains the kinematic data of 29 stroke survivors which was derived from the [27]. Participants were included when they were at least six months post-stroke, aged > 40 years, had a Mini-Mental-State-Examination (MMSE) score of at least 24 to follow instructions [28] and had a minimum functional ambulation category (FAC) of 3. The second sample contains the kinematic data of 42 healthy participants and was derived from the publicly available data set of Fukuchi et al. (approval number: CAAE: 53063315.7.0000.5594) [29]. All participants were capable of walking without handrail support during the data collection period. Further participant characteristics are displayed in Table 1.

**Table 1:** Participant characteristics of the general data set and the test split: N = number of participants, Age in years (mean ± standard deviation (std)), height in cm (mean ± std), weight in kg (mean ± std), sex and Gait speed in m/s (mean ± std).

|  |  | **N** | **Age** | **Height** | **Weight** | **Sex m/f** | **Gait speed** |
| --- | --- | --- | --- | --- | --- | --- | --- |
| **Summary**<br>**Participants** | **Stroke survivors** | 29 | 58.6 ± 11.9 | 172.0 ± 11.4 | 85.4 ± 18.3 | 13/18 | 0.7 ± 0.3 |
| | **Healthy controls young** | 24 | 27.6 ± 18.1 | 171.1 ± 10.9 | 68.4 ± 11.2 | 14/10 | 1.2 ± 0.4 |
| | **Healthy controls older** | 18 | 62.4 ± 7.4 | 161.9 ± 9.2 | 66.7 ± 10.0 | 10/8 | 1.1 ± 0.4 |
| **Summary test split** | **Stroke survivors** | 8 | 52.8 ± 10.4 | 168.1 ± 12.7 | 90.4 ± 24.4 | 7/1 | 0.9 ± 0.2 |
| | **Healthy controls young** | 10 | 28.4 ± 4.6 | 171.4 ± 11.1 | 68.8 ± 9.4 | 5/5 | 1.2 ± 0.4 |
| | **Healthy controls older** | 8 | 64.0 ± 9.6 | 164.7 ± 8.6 | 66.7 ± 9.5 | 6/2 | 1.2 ± 0.4 |

### Set-up and Procedure

Both data sets were collected during treadmill walking. The 3D trajectories of 47 retroreflective markers of the stroke survivors were captured with ten infrared cameras (100 Hz, Vicon, Vicon Motion Systems, Oxford) [27]. As for the healthy cohort, their kinematic data using the 3D trajectories of 26 retroreflective markers were collected with 12 infrared cameras (150 Hz, Raptor-4; Motion Analysis Corporation, Santa Rosa, CA, USA) [29]. Further information on the experimental setup we refer to the cited articles.

### Data pre-processing

The marker trajectories of the stroke survivors were mirrored if their paretic limb was on the left side. This ensured that the side of the impairment was not affecting the outcome of the VAE model.

Joint angles in 3D were calculated according to the ISB recommendations [30] for the hip, knee and ankle joints of both limbs. To ensure that the joint angles were not affected by either within-study sample or between-study sample differences in marker placement, the 3D joint angles of the reference pose were subtracted from the dynamic joint angles.

All data were resampled towards 50 Hz as the two data sets were acquired at different sampling frequencies.

The right foot contact events were determined using the coordinate-based treadmill algorithm [31] and were used to segment each participant’s measurement into multiple epochs of four seconds (200 frames) of data. Each epoch had a maximum of 50% overlap with the previous epoch depending on the moment of foot contact. The different joint angle trajectories were min-max normalized before training the model.

### Training and test set

In total, 5876 periods of 4 seconds formed the input data for the VAE. Each period contains the time series of the six lower-limb joints angles in three dimensions, thus an 18 by 200 input matrix. The data was split into a train and test set subject-wise to avoid data leakage, whereby the test set consisted of 2300 periods (35%). A detailed description of the train and test split is presented in Table 1.

### Models and validation

A one-dimensional convolutional variational autoencoder (VAE) [32] was used as an unsupervised deep-learning technique to analyse the gait patterns. The encoder contains 3 convolutional layers, a flatten and dense layer. The information from the encoder is represented in a defined number of latent variables, each containing a mean and standard deviation. The decoder was used to reconstruct the input based on the latent space. The decoder is symmetrical to the (supplementary material, S1). To approximate the intractable posterior, a Gaussian distribution (which consists of two trainable parameters µ and σ) is chosen as the variational inference scheme. The VAE was trained to minimize the summed reconstruction error between the input and reconstructed signal and the Kullback-Leibler divergence loss [33]. Minimizing the latter at the same time as the reconstruction error allows for a “smooth” and “regular” latent space, in which two similar vectors decode to similar signals, and in which a sampled vector decodes to a realistic signal. The model was trained on the training data set using 40 epochs. Model validation was based on the test split. The training process was stopped in case the validation loss did not improve anymore for at least 25 repetitions.

To minimize the number of latent features while preserving maximum information we developed models using 2, 3, 4 and 6 latent features. The final decision on the number of latent features was based on a minimal number of latent features without a significant reduction in model performances.

These were evaluated by determining how well the VAE was able to reconstruct the walking pattern. We calculated the root mean squared error (RMSE), RMSE normalized to the joint’s range of motion (nRMSE) and Pearson correlation coefficient r between the original and reconstructed time series. Additionally, to validate the regularity of the latent space, the time series of three randomly chosen points of the latent space were reconstructed (Figure S3).

### Identification of the individual’s gait fingerprint

Finally, the size of the area covered by each individual within the latent space was assessed. Therefore the centre of their latent features and the Euclidean distance ε of the data points to the participant’s centre were calculated:

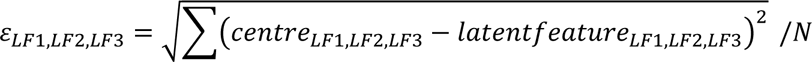

Where ε is the root mean of the sum of the squared distances normalized to the number of gait trails (N) per participant.

The size of each individual’s data cloud within the latent space was then expressed as the volume calculated from the three Euclidean distances:

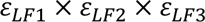

To compare the data cloud sizes between the stroke survivors and the healthy controls, the Mann-Whitney-U t-test was performed.

All procedures follow the EQUATOR network Recommendations for Reporting Machine Learning Analyses in Clinical Research [34].

## Results

### Optimising the number of latent features

While the reconstruction error was not significantly different when decoding the latent space from two, three, four or six latent features, the correlation coefficient between the reconstructed and the original signal improved when increasing the number of latent features from two to three without further improvements when increasing the number of latent features even more (*Figure 1A*). All results presented in the following are therefore based on three latent features.

**Figure 1:**
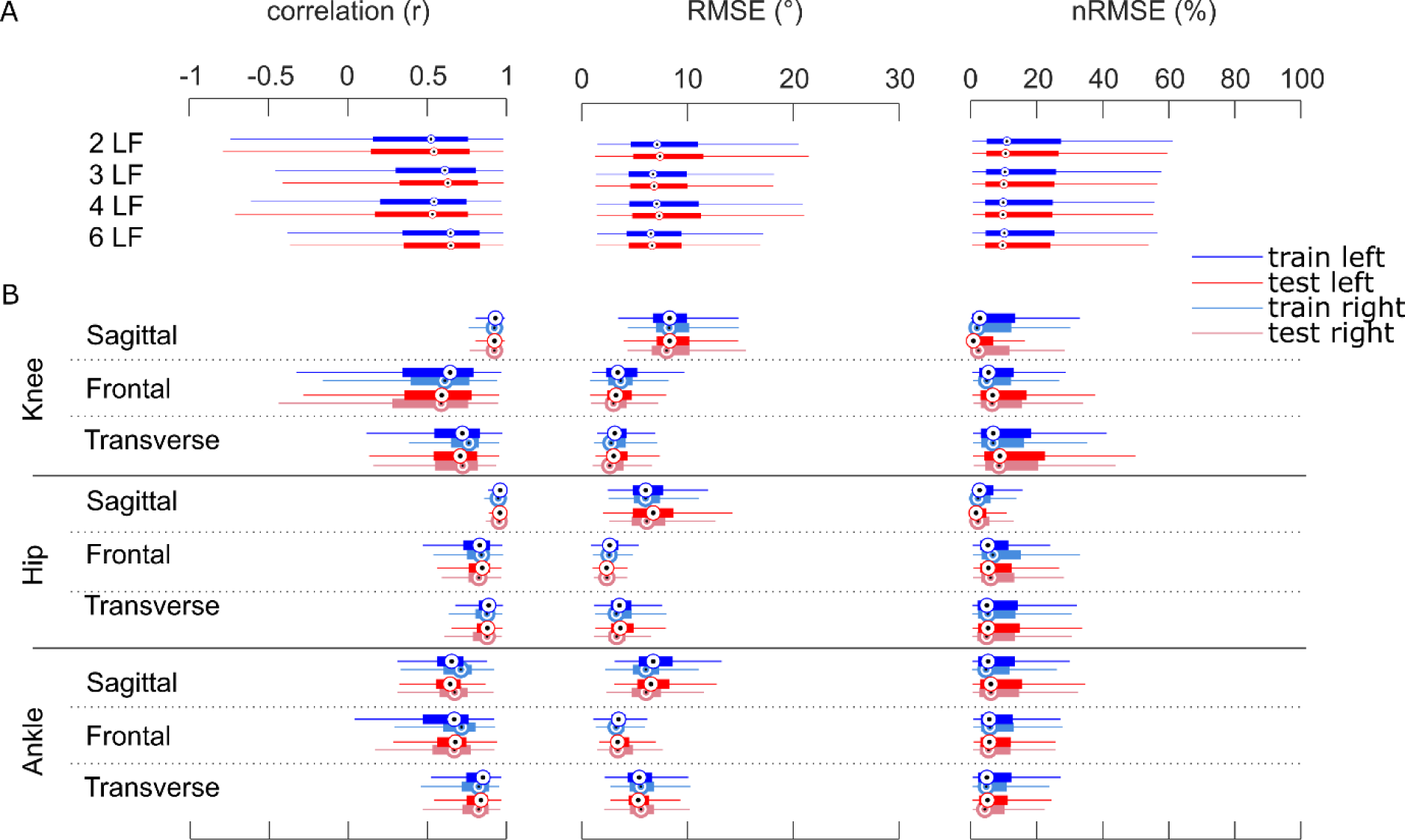
Validation results: Pearson’s r, RMSE and nRMSE for all input channels. Red boxplots refer to the reconstruction accuracy of the test set. Blue boxplots refer to the reconstruction accuracy of the train set. Circles = median, error bars = SD. Top row: comparison of model performance for two, three, four and six latent features (LF), results were averaged over all joints and motion planes. Lower rows: Model performance for three LF for the knee, hip and ankle joint angles in all motion planes. The light blue and light red boxplots indicate the reconstruction accuracy of the right (for the stroke survivors – the affected) leg, displaying only minor differences when compared to the left leg.

### Evaluation of VAE model

To assess the ability of the network to reduce the gait data to three latent features with minimal loss of relevant information, the time series of the training and the test set were reconstructed from the latent features. The error between the reconstructed and the raw input data is displayed in *Figure 1B*. The RMSE and the nRMSE for the test and the train set showed comparable results (RMSE: 6.03° ± 2.71 and 5.76° ± 2.68, nRMSE: 23.02% ± 4.05 and 23.36% ± 4.13). Due to the higher variability relative to the range of motion, the nRMSE was higher for the non-sagittal movements while the RMSE was highest for the knee flexion angle (11.70° ± 0.23 averaged over training and test set).

The correlation between the reconstructed and the raw data showed a high coefficient (range 0.70 – 0.91) for the sagittal knee and all motion planes of the left and right hip joint angles (*Figure 1B*). A moderate relationship (range 0.52 – 0.69) for all other joints and planes except the right frontal knee joint angle which only showed a low to moderate correlation (r = 0.40).

The reconstruction accuracy was not systematically different between the healthy cohort and the stroke survivors (Figure S2), which indicated that the network is capable of learning both healthy and pathologic, potentially more inconsistent gait patterns. There was no difference between the left and right leg’s joint angles (*Figure 1B*). Further, the reconstruction of three arbitrary points from the latent space resulted in realistic gait patterns (Figure S3).

### Gait fingerprint

The quality of the network was assessed in its ability to preserve relevant information which resulted in the successful discrimination between the gait pattern of stroke survivors and the healthy controls. Figure 2A displays the encoded data of the test set. The stroke survivors manifest into three main groups, located on the outer rim of the healthy controls.

**Figure 2:**
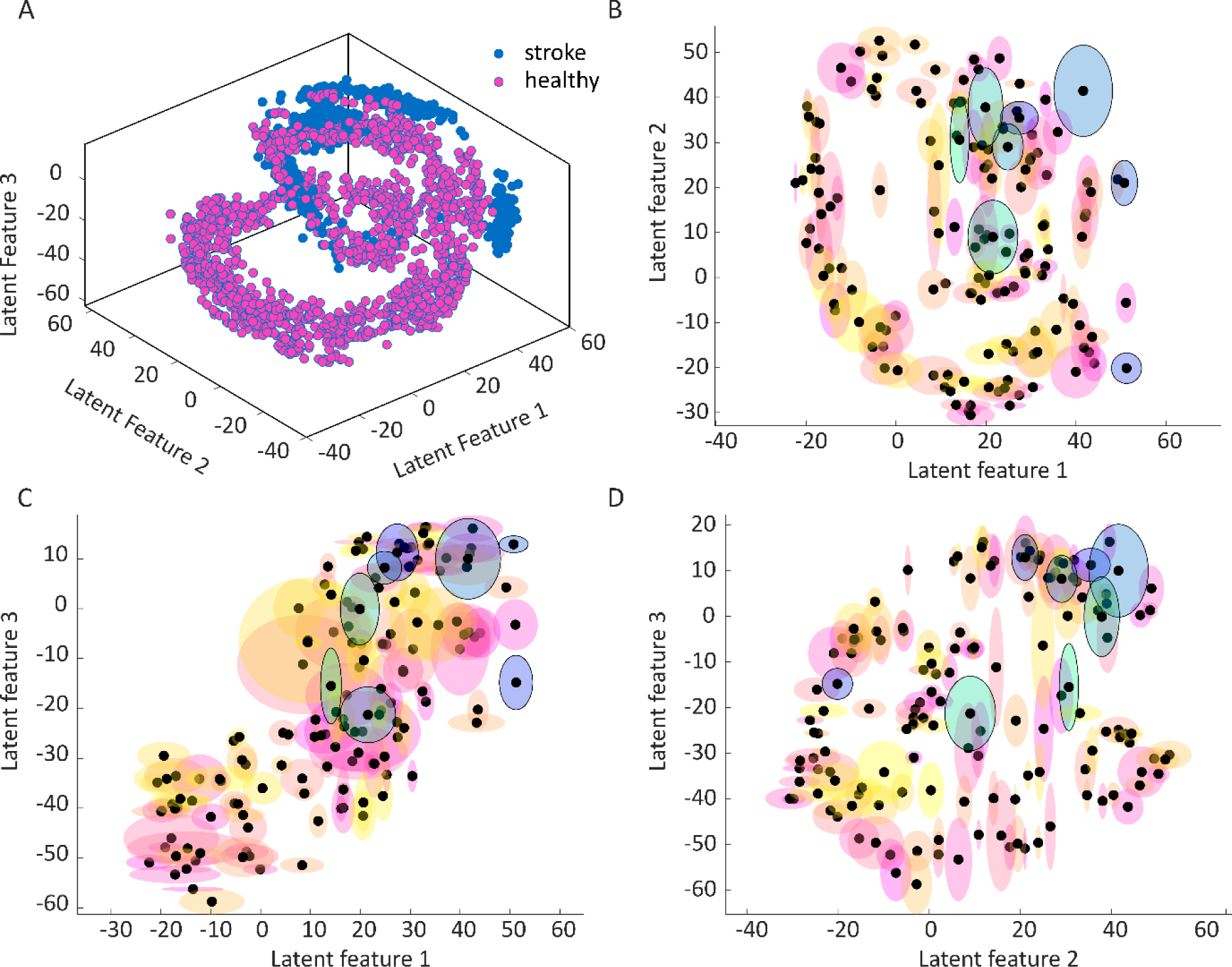
A: The stroke survivor’s data (blue) is located on the outer rim of the healthy cohort’s data (pink). For interactive figures visit: https://mybinder.org/v2/gh/SinaDavid/VAE/main?urlpath=%2Ftree%2F B-D: Representation of the Euclidean distances and centres of the clusters of each included individual, the black dots represent the centre of each participant, while the shaded areas represent the space covered by their data. The ellipsoids are based on the Euclidean distance in the 3 directions of the latent space. The areas, represented in blue/green are representing the stroke survivors, expressing the large variability within their gait pattern. The areas coloured in red/yellow represent the healthy controls.

The covered space of the included stroke patients, represented as the 3D Euclidean distance from the participant’s centre is significantly higher than those of the healthy controls for all latent features except LF2 (p<0.001, see Table 2). Also, the stroke survivors’ volume of the data cloud was significantly larger (p<0.001), expressing that stroke gait pattern is less uniform.

**Table 2:** Covered area of the individual data clouds within the latent space represented as the Euclidean distances ε of the three latent features (LF) and the resultant size. The statistical outcome of the Mann-Whitney U test is represented by the p-value.

|  | Stroke survivors | Healthy controls | p-value |
| --- | --- | --- | --- |
| $\varepsilon$ LF 1 (mean ± std) | $4.48 \pm 3.32$ | $2.61 \pm 1.49$ | <0.001 |
| $\varepsilon$ LF 2 (mean ± std) | $6.09 \pm 4.89$ | $4.16 \pm 3.15$ | 0.059 |
| $\varepsilon$ LF 3 (mean ± std) | $6.06 \pm 4.08$ | $3.02 \pm 2.41$ | <0.001 |
| Size of data cloud (mean ± std) | $266.84 \pm 375.71$ | $48.97 \pm 125.65$ | <0.001 |

## Discussion

This study aimed to determine if a VAE can describe gait data with a limited number of features while preserving all relevant information, free from theory-driven constraints. It was hypothesized that a VAE is capable of recognizing gait patterns of healthy and pathologic gait. For this purpose, the data set of stroke survivors and a healthy population were merged, and their 3D lower-limb joint angles were used as the input for the VAE, which encoded the input into three latent features. These latent features were used as the 3D coordinates to map each of the gait segments in the 3D space to further analyse the specific location within this space.

The most promising result of this study was that the VAE recognizes distinct characteristics of gait patterns and locates them in different regions of the latent space (Figure 3). Most participants covered only a small area, which indicates that the network indeed recognized a participant-specific gait pattern distinguishing the individual from others. However, some participants covered larger areas, expressed also in larger Euclidean distances from the centre of their data (Figure 2B-D) and an overall larger data cloud. The stroke survivors’ areas were significantly larger, which is not a surprising result, as they are reported to have less uniform gait patterns than healthy individuals [16]. We propose that the centre and the area covered by one’s data cloud within the latent space as an easy-to-report measure of this gait pattern to be monitored throughout gait training. Variability in gait is a well-discussed parameter, usually assessed using entropy, standard deviation or similar approaches. While the standard deviation as a linear measure is not a good representation of a non-linear data set [11], entropy is, but its interpretation is difficult. Both, the location and the size of the area are clear representatives of the individual’s gait fingerprint. The size of the data cloud is the direct outcome of the variability, and a decrease represents a more uniform gait pattern. However, using the size of a patient’s data cloud as a variability measure is just a side product of the VAE. A network that is capable of learning complex information, can be utilised in several ways, not just for variability estimation.

**Figure 3:**
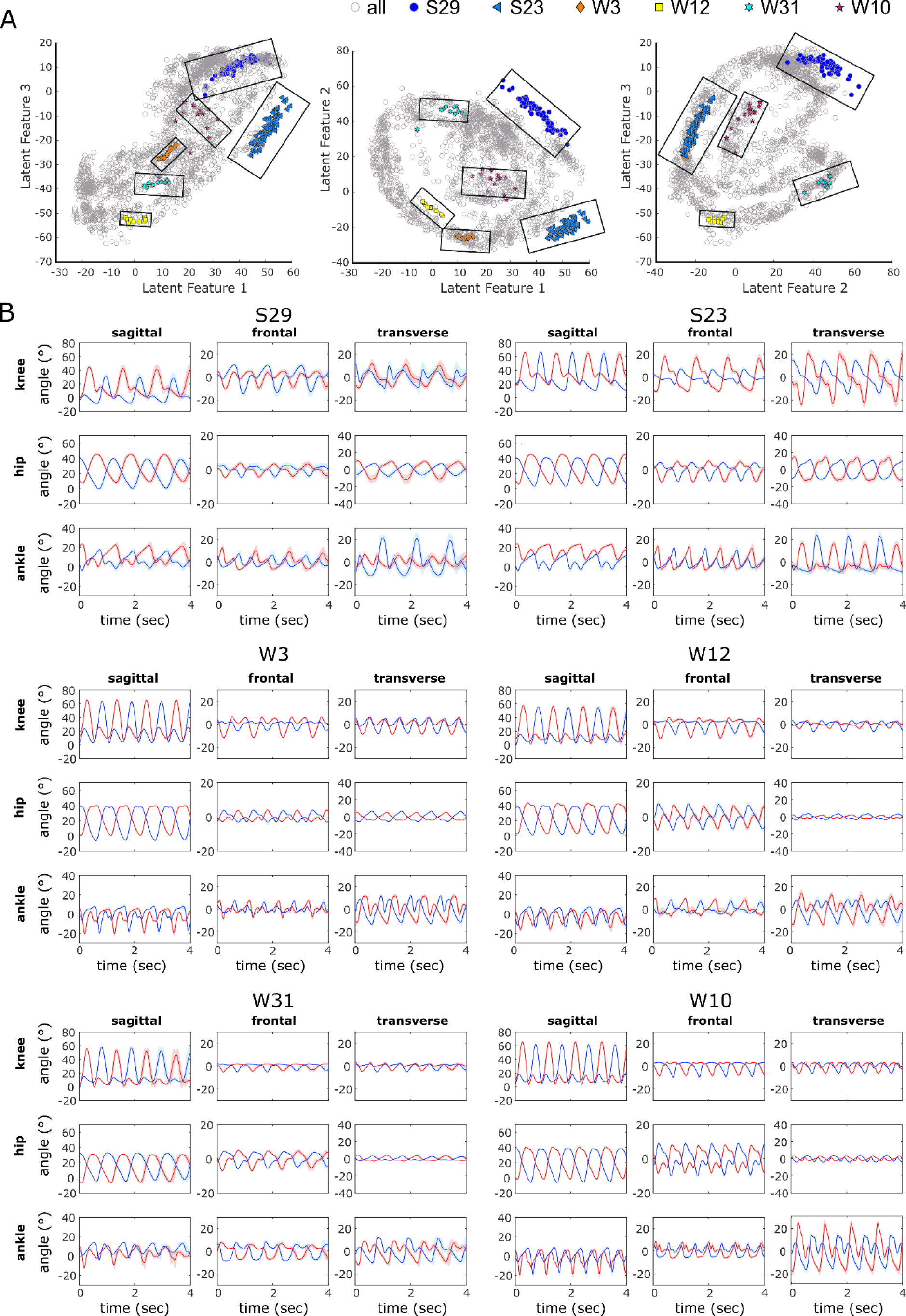
Regional analysis of six selected participants. A) representation of the latent features of each of the gait trials. B) mean (bold line) and standard deviation (shaded area) of the reconstruction of the joint angles of six selected participants from their three latent features. Blue: right leg, red: left leg. Participants S16, S19 and S21 are from the group of stroke survivors while the other three participants are from the healthy cohort.

As all lower limb joint angles were used to train the VAE, the location of a single patient within the latent space represents wholesome information about the gait pattern. This location is not biased by any hypothesis, it is data-driven. Further, the transition of location is continuous, which is advantageous as also small changes in the gait pattern are likely to be recognized. This makes the use of VAEs superior to classification methods where the features are forced into groups which neglects the continuity of changes. This opens a new opportunity to assess the gait quality of a patient. Simply speaking, the location of the patient on the map relative to e.g. a healthy cohort can be used as a starting point for the rehabilitation process. By knowing the characteristics of a certain region of the map, the joint and plane of highest priority can be defined and prioritized during rehabilitation. During treatment, the patient can be monitored by presenting the new gait data to the VAE and tracking the location within the latent space. With this, the clinician’s assessment of the rehabilitation can be supported in an objective, efficient and wholesome manner.

The quality of the used VAE was high enough to satisfyingly reduce the high dimensional gait data towards three latent features while still being able to distinguish between individuals. This is reflected by high Pearson correlation coefficients and low RMSE and nRMSE results. However, it has to be mentioned that the reconstruction of the non-sagittal plane joint angles was less accurate than the sagittal plane movement. This is not a specific issue of VAEs but has also been reported for supervised learning algorithms – resulting from the combination of low movement amplitude and high movement variability [35–37]. Therefore, it is more difficult for any algorithm to learn the pattern. To extract individual gait patterns, high reconstruction accuracy is preferred, but the current accuracy is sufficient since the joint angles aren’t used for decision-making. Increasing the dataset could further improve accuracy. The outer edge of the latent space had lower reconstruction accuracy than the centre, as it represents the gait patterns of the slowest and fastest walkers, trained on limited data. To extend the area of high accuracy, the network could be trained with more data from noisy conditions or more impaired gait patterns.

The latent space forms a spiral structure, contrary to the ideal of finding completely disentangled latent features where changes in one unit affect only one generative factor [38]. In this case, all three latent features together describe the data variation. It’s noted that improving disentanglement often increases reconstruction loss [38,39]. Future research should explore whether a different balance between entanglement and reconstruction loss could enhance the research outcomes.

The presented results are explorative. To be applicable in clinical practice, it is necessary to increase the sample size of the training set especially with data coming from pathologic gait. Having data sets of patients that were collected in comparable environments was restricted in the past, however, this will change in the future. In recent years, the number of publicly available data sets increased. With many initiatives promoting open science, this process should become the new normal soon, enabling us to reach the aim of increasing sample size.

Optical motion capture data, like in this study, is often impractical in clinical settings due to high costs and time requirements. Future efforts will focus on using data from wearable sensors or 2D video cameras, which are more affordable and user-friendly.

## Conclusion

The results demonstrate that deep unsupervised learning algorithms effectively assess individual movement patterns. VAEs can detect subtle gait differences, providing objective and data-driven evaluations of patient gait alterations. Representing complex data in a 3D map enables clinicians to assess gait without predefined assumptions, while monitoring extracted features helps track rehabilitation progress. This approach enhances clinical efficiency and improves patient outcomes through personalized therapy.

## Data Availability

All data produced in the present study are available upon reasonable request to the authors.

## Acknowledgement.

The authors would like to thank the research group of Fukuchi et al. for making their data publicly available.

## Author contributions

SD wrote the manuscript and prepared the data for analysis and postprocessing of the results. MP collected the stroke data set, was responsible for generating the network architecture and edited the manuscript. SD and MP were responsible for interpreting the results. SG, CG and YL consulted and optimized the network architecture and edited the manuscript.

## Data availability statement

The initialized and trained weights together with the network code are available at: https://github.com/SinaDavid/VAE.

## References

[1] W. Yu, H. Yu, Y. Huang, L. Wang, Generalized Inter-class Loss for Gait Recognition, (2022). 10.1145/3503161.3548311.

[2] D. Pinčić, D. Sušanj, K. Lenac, Gait Recognition with Self-Supervised Learning of Gait Features Based on Vision Transformers, Sensors 22 (2022) 7140. 10.3390/s22197140.

[3] K.A. Duncanson, S. Thwaites, D. Booth, G. Hanly, W.S.P. Robertson, E. Abbasnejad, D. Thewlis, Deep Metric Learning for Scalable Gait-Based Person Re-Identification Using Force Platform Data, Sensors 23 (2023) 3392. 10.3390/s23073392.

[4] H.-S. Kim, S.-C. Chung, M.-H. Choi, S.-Y. Gim, W.-R. Kim, G.-R. Tack, D.-W. Lim, S.-K. Chun, J.-W. Kim, K.-R. Mun, Primary and secondary gait deviations of stroke survivors and their association with gait performance, J Phys Ther Sci 28 (2016) 2634–2640. 10.1589/jpts.28.2634.

[5] J.W. Gersten, W. Orr, External work of walking in hemiparetic patients., Scand J Rehabil Med 3 (1971) 85–8.

[6] P.J. Corcoran, R.H. Jebsen, G.L. Brengelmann, B.C. Simons, Effects of plastic and metal leg braces on speed and energy cost of hemiparetic ambulation., Arch Phys Med Rehabil 51 (1970) 69–77.

[7] C.B. de Oliveira, Í.R.T. de Medeiros, N.A. Ferreira, M.E. Greters, A.B. Conforto, Balance control in hemiparetic stroke patients: main tools for evaluation., J Rehabil Res Dev 45 (2008). 10.1682/JRRD.2007.09.0150.

[8] J. Park, T.-H. Kim, The effects of balance and gait function on quality of life of stroke patients, NeuroRehabilitation 44 (2019) 37–41. 10.3233/NRE-182467.

[9] S. Nadeau, M. Betschart, F. Bethoux, Gait Analysis for Poststroke Rehabilitation, Phys Med Rehabil Clin N Am 24 (2013) 265–276. 10.1016/j.pmr.2012.11.007.

[10] D. Levine, J. Richards, M.W. Whittle, Whittle’s gait analysis, Elsevier health sciences, 2012.

[11] R.T. Harbourne, N. Stergiou, Movement Variability and the Use of Nonlinear Tools: Principles to Guide Physical Therapist Practice, Phys Ther 89 (2009) 267–282. 10.2522/ptj.20080130.

[12] D.L. Skaggs, S.A. Rethlefsen, R.M. Kay, S.W. Dennis, R.A.K. Reynolds, V.T. Tolo, Variability in gait analysis interpretation, Journal of Pediatric Orthopaedics 20 (2000) 759–764.

[13] B. Toro, C. Nester, P. Farren, A review of observational gait assessment in clinical practice, Physiother Theory Pract 19 (2003) 137–149. 10.1080/09593980307964.

[14] D.M. Mohan, A.H. Khandoker, S.A. Wasti, S. Ismail Ibrahim Ismail Alali, H.F. Jelinek, K. Khalaf, Assessment Methods of Post-stroke Gait: A Scoping Review of Technology-Driven Approaches to Gait Characterization and Analysis, Front Neurol 12 (2021). 10.3389/fneur.2021.650024.

[15] H. Lau, K. Tong, H. Zhu, Support vector machine for classification of walking conditions of persons after stroke with dropped foot, Hum Mov Sci 28 (2009) 504–514. 10.1016/j.humov.2008.12.003.

[16] M. Punt, S.M. Bruijn, K.S. van Schooten, M. Pijnappels, I.G. van de Port, H. Wittink, J.H. van Dieën, Characteristics of daily life gait in fall and non fall-prone stroke survivors and controls, J Neuroeng Rehabil 13 (2016) 67. 10.1186/s12984-016-0176-z.

[17] C. Adans-Dester, N. Hankov, A. O’Brien, G. Vergara-Diaz, R. Black-Schaffer, R. Zafonte, J. Dy, S.I. Lee, P. Bonato, Enabling precision rehabilitation interventions using wearable sensors and machine learning to track motor recovery, NPJ Digit Med 3 (2020) 121. 10.1038/s41746-020-00328-w.

[18] S. Sapienza, C. Adans-Dester, A. OBrien, G. Vergara-Diaz, S. Lee, S. Patel, R. Black-Schaffer, R. Zafonte, P. Bonato, C. Meagher, A.-M. Hughes, J. Burridge, D. Demarchi, Using a Minimum Set of Wearable Sensors to Assess Quality of Movement in Stroke Survivors, in: 2017 IEEE/ACM International Conference on Connected Health: Applications, Systems and Engineering Technologies (CHASE), IEEE, 2017: pp. 284–285. 10.1109/CHASE.2017.104.

[19] K. Kaczmarczyk, A. Wit, M. Krawczyk, J. Zaborski, Gait classification in post-stroke patients using artificial neural networks, Gait Posture 30 (2009) 207–210. 10.1016/j.gaitpost.2009.04.010.

[20] S. Mulroy, J. Gronley, W. Weiss, C. Newsam, J. Perry, Use of cluster analysis for gait pattern classification of patients in the early and late recovery phases following stroke, Gait Posture 18 (2003) 114–125. 10.1016/S0966-6362(02)00165-0.

[21] N. Razfar, R. Kashef, F. Mohammadi, Automatic Post-Stroke Severity Assessment Using Novel Unsupervised Consensus Learning for Wearable and Camera-Based Sensor Datasets, Sensors 23 (2023) 5513. 10.3390/s23125513.

[22] P. Krondorfer, D. Slijepčević, F. Unglaube, A. Kranzl, C. Breiteneder, M. Zeppelzauer, B. Horsak, Deep learning-based similarity retrieval in clinical 3d gait analysis, Gait Posture 90 (2021) 127–128.

[23] F. Horst, D. Slijepcevic, M. Simak, B. Horsak, W.I. Schöllhorn, M. Zeppelzauer, Modeling biological individuality using machine learning: A study on human gait, Comput Struct Biotechnol J 21 (2023) 3414–3423. 10.1016/j.csbj.2023.06.009.

[24] D.P. Kingma, M. Welling, An Introduction to Variational Autoencoders, Foundations and Trends® in Machine Learning 12 (2019). 10.1561/2200000056.

[25] N. Simidjievski, C. Bodnar, I. Tariq, P. Scherer, H. Andres Terre, Z. Shams, M. Jamnik, P. Liò, Variational Autoencoders for Cancer Data Integration: Design Principles and Computational Practice, Front Genet 10 (2019). 10.3389/fgene.2019.01205.

[26] C.K. Balasubramanian, R.R. Neptune, S.A. Kautz, Variability in spatiotemporal step characteristics and its relationship to walking performance post-stroke, Gait Posture 29 (2009) 408–414. 10.1016/j.gaitpost.2008.10.061.

[27] M. Punt, S.M. Bruijn, S. Roeles, I.G. van de Port, H. Wittink, J.H. van Dieën, Responses to gait perturbations in stroke survivors who prospectively experienced falls or no falls, J Biomech 55 (2017). 10.1016/j.jbiomech.2017.02.010.

[28] M.F. Folstein, The Mini-Mental State Examination, Arch Gen Psychiatry 40 (1983) 812. 10.1001/archpsyc.1983.01790060110016.

[29] C.A. Fukuchi, R.K. Fukuchi, M. Duarte, A public dataset of overground and treadmill walking kinematics and kinetics in healthy individuals., PeerJ 6 (2018). 10.7717/peerj.4640.

[30] G. Wu, S. Siegler, P. Allard, C. Kirtley, A. Leardini, D. Rosenbaum, M. Whittle, D.D. D’Lima, L. Cristofolini, H. Witte, O. Schmid, I. Stokes, ISB recommendation on definitions of joint coordinate system of various joints for the reporting of human joint motion—part I, J Biomech 35 (2002) 543–548. 10.1016/S0021-9290(01)00222-6.

[31] J.A. Zeni, J.G. Richards, J.S. Higginson, Two simple methods for determining gait events during treadmill and overground walking using kinematic data, Gait Posture 27 (2008) 710–714. 10.1016/j.gaitpost.2007.07.007.

[32] D.P. Kingma, M. Welling, Auto-Encoding Variational Bayes, (2013).

[33] I. Csiszar, I-Divergence Geometry of Probability Distributions and Minimization Problems, The Annals of Probability 3 (1975). 10.1214/aop/1176996454.

[34] L.M. Stevens, B.J. Mortazavi, R.C. Deo, L. Curtis, D.P. Kao, Recommendations for Reporting Machine Learning Analyses in Clinical Research, Circ Cardiovasc Qual Outcomes 13 (2020). 10.1161/CIRCOUTCOMES.120.006556.

[35] M. Mundt, A. Koeppe, F. Bamer, S. David, B. Markert, Artificial Neural Networks in Motion Analysis-Applications of Unsupervised and Heuristic Feature Selection Techniques., Sensors (Basel) 20 (2020). 10.3390/s20164581.

[36] J. Lebleu, T. Gosseye, C. Detrembleur, P. Mahaudens, O. Cartiaux, M. Penta, Lower Limb Kinematics Using Inertial Sensors during Locomotion: Accuracy and Reproducibility of Joint Angle Calculations with Different Sensor-to-Segment Calibrations, Sensors 20 (2020) 715. 10.3390/s20030715.

[37] R.D. Gurchiek, N. Cheney, R.S. McGinnis, Estimating Biomechanical Time-Series with Wearable Sensors: A Systematic Review of Machine Learning Techniques, Sensors 19 (2019) 5227. 10.3390/s19235227.

[38] M.D. Hoffman, M.J. Johnson, Elbo surgery: yet another way to carve up the variational evidence lower bound, in: Workshop in Advances in Approximate Bayesian Inference, NIPS, 2016.

[39] I. Higgins, L. Matthey, A. Pal, C. Burgess, X. Glorot, M. Botvinick, S. Mohamed, A. Lerchner, beta-vae: Learning basic visual concepts with a constrained variational framework, (2016).

