## Supplementary Material for "A novel approach to identify the fingerprint of stroke gait using deep unsupervised learning"

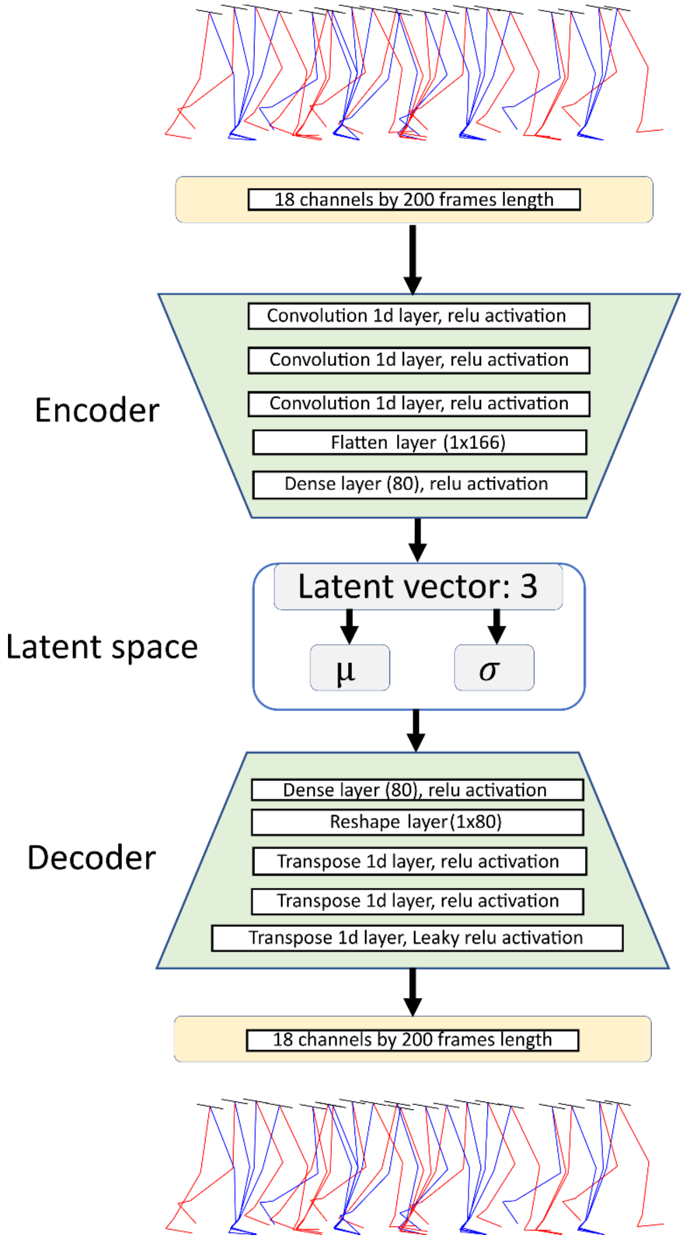


*Figure S1: Network architecture of the variational autoencoder (VAE)*

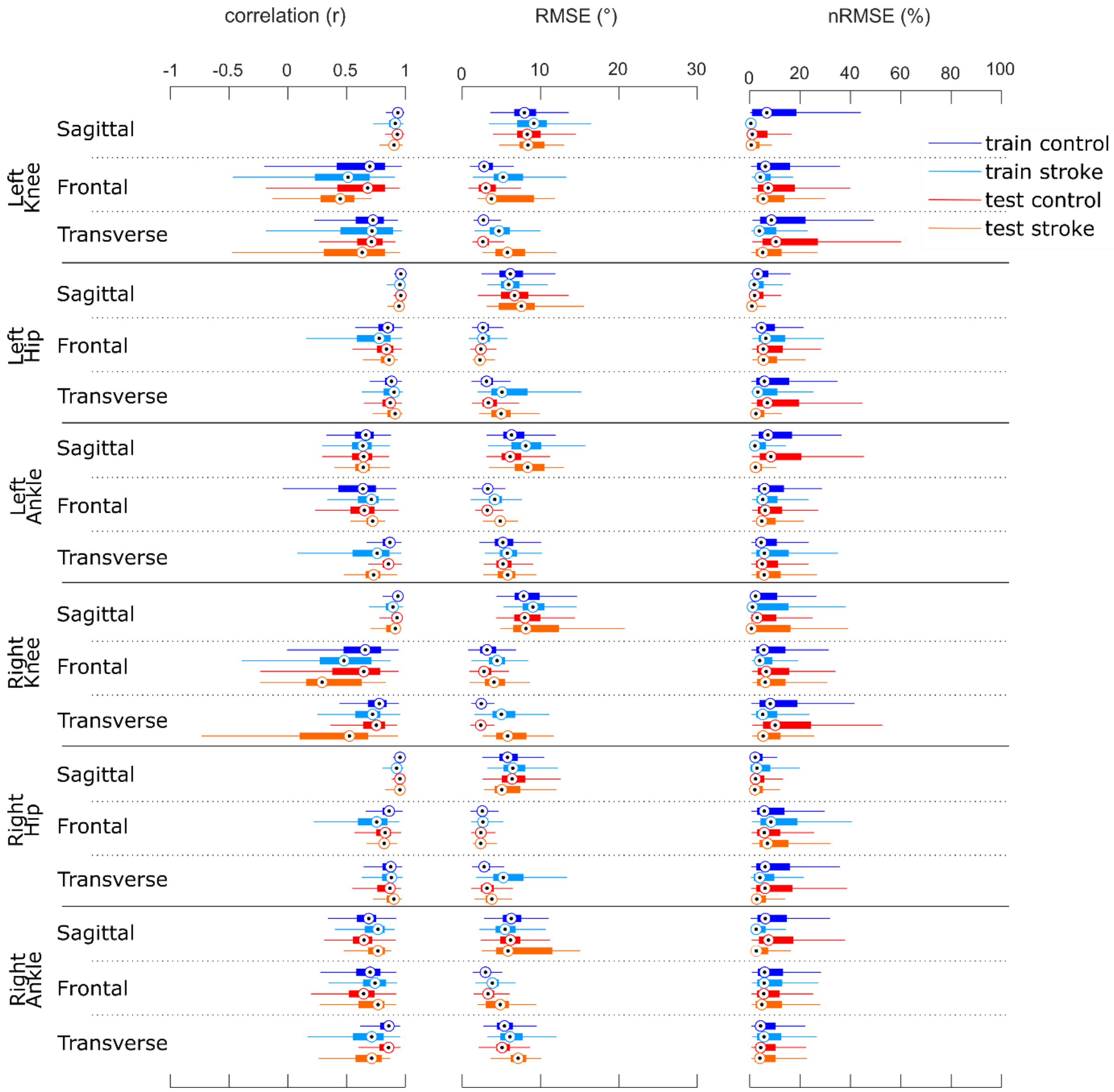


*Figure S2: Validation results: Pearson's r, RMSE and nRMSE for all input channels. Red boxplots refer to the reconstruction accuracy of the test set. Blue boxplots refer to the reconstruction accuracy of the train set. Light blue and light red refer to the data of the stroke survivors, and dark blue and dark red represent the results of the healthy controls.*

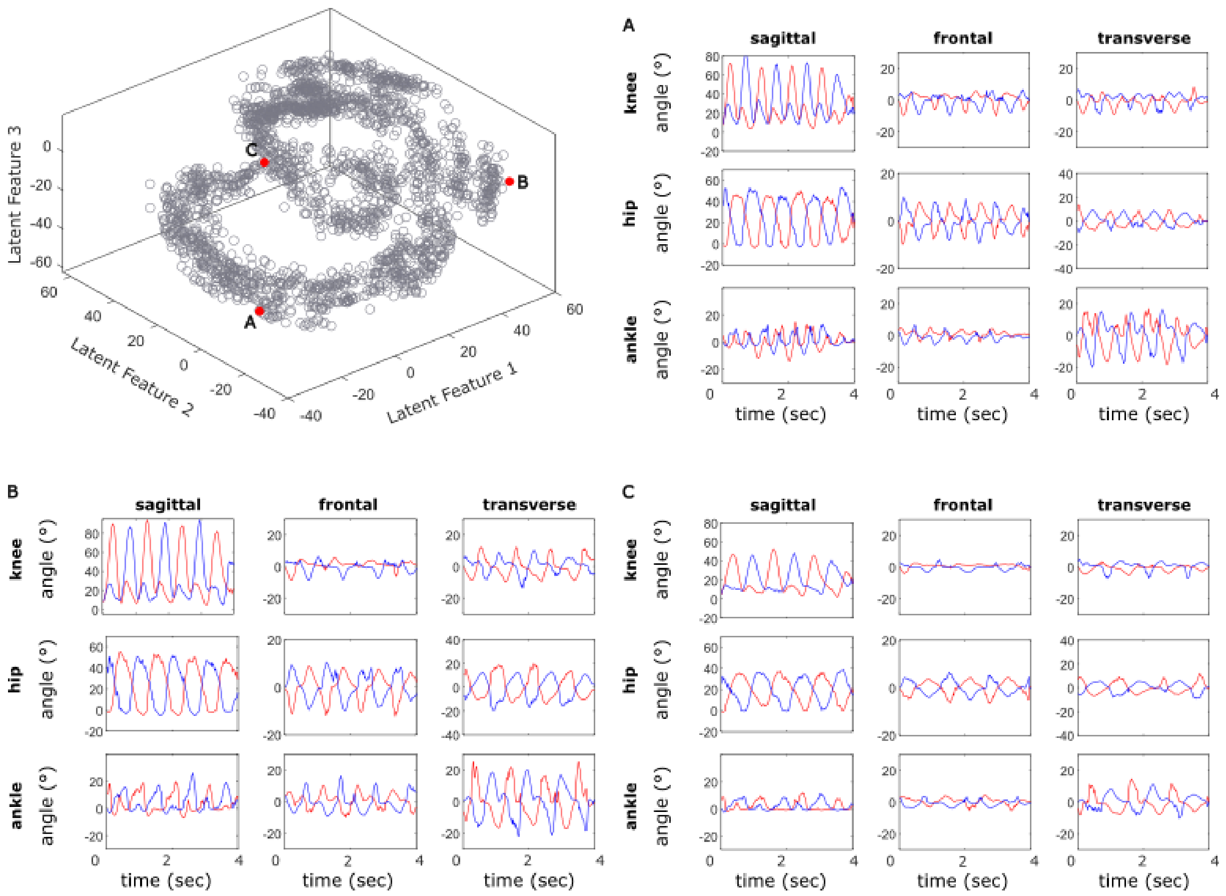


*Figure S3: To validate the regularity of the latent space, the time series of 3 randomly chosen points (A-C) from the latent space were reconstructed and show joint angles within a normal range. Blue: right leg, red: left leg.*
